# One-shot immunization with Sputnik Light (the first component of Sputnik V vaccine) is effective against SARS-CoV-2 Delta variant: efficacy data on the use of the vaccine in civil circulation in Moscow

**DOI:** 10.1101/2021.10.08.21264715

**Authors:** Inna V Dolzhikova, Vladimir A Gushchin, Dmitry V Shcheblyakov, Alexander N. Tsybin, Alexey M Shchetinin, Andrey A Pochtovyi, Andrey B Komissarov, Denis A. Kleymenov, Nadezhda A Kuznetsova, Amir I Tukhvatulin, Olga V Zubkova, Alina S Dzharullaeva, Anna V Kovyrshina, Nadezhda L Lubenets, Daria M Grousova, Alina S Erokhova, Fatima M Izhaeva, Olga Popova, Tatiana A Ozharovskaya, Alexander S Semikhin, Elizaveta A Tokarskaya, Maksim M Shmarov, Natalia A Nikitenko, Elena V Shidlovskaya, Evgeniia N. Bykonia, Elena P. Mazunina, Elizaveta V Divisenko, Lyudmila A Vasilchenko, Liubov I. Popova, Daria M Danilenko, Dmitry A Lioznov, Artem P Tkachuk, Sergei V Borisevich, Boris S Naroditsky, Denis Y Logunov, Alexander L Gintsburg

## Abstract

**Objectives:** Vaccination remains the most effective response to the COVID-19 pandemic. Most vaccines use two-dose regimens. In turn, single-dose vaccines also have high potential, since, on the one hand, they simplify the vaccination program, make it more accessible and convenient for more people around the world, and on the other hand, they are better suited for subsequent revaccination. However, there is not enough data on the effectiveness of single-dose vaccine variants against new genetic lines to assess their current potential. It is not clear how much a single dose of immunization protects against the globally dominant delta variant. In this work, we investigated the effectiveness of a single dose vaccine (Sputnik Light, the first component of Sputnik V vaccine) against the Delta variant in Moscow.

**Methods:** To assess the effectiveness of one dose of viral vector vaccine based on rAd26 against the delta variant in Moscow, we used data from the Moscow registries of vaccination against COVID-19 and the incidence of COVID-19. The availability of data on the number of seropositive residents of Moscow made it possible to consider the size of the immune layer formed because of a previous COVID-19 disease or vaccination. To calculate the effectiveness, the proportion of COVID-19 cases among those vaccinated with a single dose and the proportion of cases among those who were not vaccinated in July 2021.

**Results:** Our data indicate that throughout July 2021, the dominant variant of the coronavirus at the level of 99.5% in Moscow was the SARS-CoV-2 delta variant and its subsidiary lines. Considering the immune layer of 46% allowed us to calculate the effectiveness of a one-shot vaccine against the delta variant in Moscow during the first three months after vaccination at the level of 69.85% (95% confidence interval [CI], 64.08 to 74.70). In the 18-29-year-old group, the overall vaccine efficacy against the delta variant was 88.61%, in the 18-59 group - 75.28%. Sputnik Light demonstrates higher efficacy against Delta variant than many two-shot vaccines.

**Conclusion:** The results indicate a high efficacy of a single immunization first component of Sputnik V vaccine against delta variant among young and middle-aged people, at least during the first 3 months after receiving the one-shot vaccine.

## Introduction

Today, there is a fairly large selection of vaccines for the prevention of COVID-19, which have shown high efficiency at the stage of clinical trials. [1-4]. However, the efficacy against the SARS-CoV-2 delta variant for many vaccines has decreased [5-6], which explains the increase in the global incidence of COVID-19 even against the high percentage of vaccinated in developed countries [7].

Genetic line B.1.617.2 delta was first identified in samples from patients from India [8]. This genetic line has the status of a variant of concern (VOC-21APR-02) and has a few key mutations in the S protein, including: T19R G142D E156G F157del R158del L452R T478K D614G P681R D950N. A new set of RBD mutations of the delta variant significantly reduces the virus neutralizing properties of antibodies formed by vaccination or a previous disease, promotes the formation of syncytia, allowing additional escape from antibody protection [9]. Breaking through preexisting immunity allows the virus to circulate among vaccinated and person with COVID-19 in anamnesis. Transmission of the virus among vaccinated and previously infected individuals poses a significant risk of completely losing the effectiveness of vaccines in preventing transmission of the virus and reducing mortality from COVID-19 [10].

Previously, we have developed, and tested COVID-19 vaccines based on recombinant adenoviruses (rAd): Sputnik V and Sputnik Light. The principle of action of vaccines lays in the use of adenoviral vectors that are not capable of replication in the human body but are capable of delivering the gene for the SARS-CoV-2 glycoprotein S into cells. For the Sputnik V vaccine, heterologous prime-boost immunization regimen is used when two different rAd serotypes are used: rAd26 and rAd5 [11]. In turn, the Sputnik Light vaccine is a one-shot vaccine - the first component based on rAd 26 [12]. The protective efficacy of the Sputnik V vaccine, according to the third phase of clinical trials conducted on the territory of the Russian Federation, was 91.6% [4]. Available data from studies of the effectiveness of two doses of vaccination with Sputnik V indicate that the vaccine against the delta variant is above 80% [13-14]. Data on COVID-19 cases in June 2021 shows that the effectiveness of the Sputnik V vaccine against the delta variant is about 83% [15].

It is important to understand how effective a single vaccination is. Given that the single-dose vaccine has many advantages, including a simplified vaccination program that makes it more accessible and convenient for more people around the world, there is an urgent need to obtain data on the effectiveness of the first component of Sputnik V specifically in relation to SARS-CoV-2 delta variant. In this study, we performed a preliminary calculation of the effectiveness of a single immunization with first component of Sputnik V against delta according to the Moscow vaccination registries and the incidence of COVID-19.

## Results

According to the Russian consortium for coronavirus genome sequencing – CORGI (https://corgi.center/en/) in June 2021, the frequency of the genetic line delta B.1.617.2 and its descendant lines in Russia was more than 90%, while in July 2021 it exceeded 98.5% [16]. We performed monitoring of circulating lines, analyzed 655 complete genomes of the virus in June 2021 and 654 complete genomes in July 2021 from Moscow patients and showed that the proportion of the genetic line delta B.1.617.2 and descendant lines was 93.4% and 99.5%, respectively (Supplementary Table S1). It should be noted that it apparently took no more than three months to occupy the dominant position of the delta genetic line, so if in April 2021 the proportion of the delta variant was about 2.5%, in July 2021 it was already 99.5% (Fig. 1).

**Figure 1.**
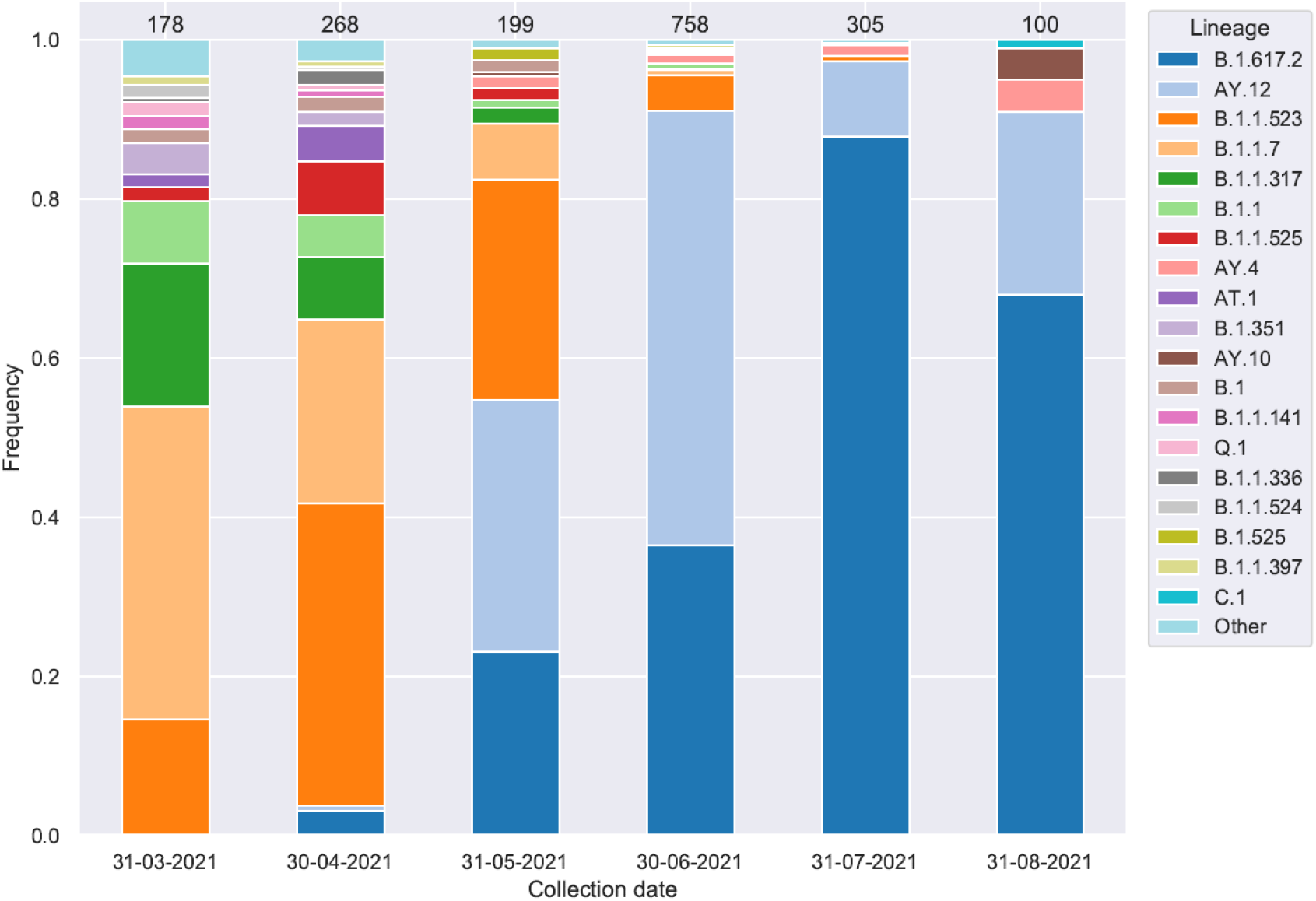
Frequency of occurrence of genetic lines in Moscow, preceding the complete displacement of all other lines by delta variant.

Among the delta’s descendant lines during the monitoring period in Moscow were B.1.617.2, AY.4, AY.5, AY.6, AY.10, AY.12, AY.20, AY.23, AY.24, but in they were mainly B.1.617.2 and AY.12. Thus, our results suggest that the calculation of vaccine efficacy in July 2021 will reflect efficacy against SARS-CoV-2 delta variant. In this regard, the July was chosen as a reference month for assessing the effectiveness of the vaccine.

We obtained data from registries of COVID-19 cases and vaccinated persons, as well as data on the number of Moscow residents in different age cohorts. Among those vaccinated with a single dose, the number of those who received the first dose of the vaccine in the interval from 21 days to 3 months after vaccination at the beginning of July and the number of cases among them during July were taken into account. In the case of non-immune people, the number of people not included in the immune layer (who did not receive any of the available vaccines, did not get sick, did not have specific antibodies at the beginning of July, ∼ 54% of the Moscow population) and the number of people who were sick among them were taken into account. The effectiveness of the vaccine was calculated in relation to the odds ratio among the group of once vaccinated and the group of non-immune, according to the WHO recommendations [17].

We found that the overall efficacy of single-dose vaccination against the delta variant was 69.85% (table 1, figure 2). In the group of 18-29 yo, the effectiveness for the vaccine was 88.61%, in the group 18-59 yo - 75.28%, while with age, the effectiveness of the vaccine tended to decrease and in the group over 60 it was 51.98% (table 1, figure 2).

**Table 1.** Efficacy of a single-dose vaccine in different groups

| | Vaccinated | | Non immune (no vaccine, no prior infection) | | p-value | VE through odds ratio<br>$VE=(1-OR)*100$ | | |
| --- | --- | --- | --- | --- | --- | --- | --- | --- |
|  | Disease | No disease | Disease | No disease |  | VE, % | 95% CI |  |
| 18+, total | 126 | 28 322 | 82 196 | 5 569 885 | <0,0001 | **69,85** | 64,08 | 74,70 |
| 18-29 | 10 | 3 926 | 15 484 | 692 374 | <0,0001 | 88,61 | 78,81 | 93,88 |
| 30-39 | 27 | 6 152 | 19 732 | 1 193 746 | <0,0001 | 73,45 | 61,24 | 81,81 |
| 40-49 | 26 | 5 957 | 14 194 | 1 061 843 | <0,0001 | 67,35 | 51,99 | 77,80 |
| 50-59 | 18 | 4 620 | 12 132 | 932 002 | <0,0001 | 70,07 | 52,44 | 81,17 |
| 60-69 | 22 | 3 599 | 10 514 | 858 207 | 0,0004 | 50,10 | 24,09 | 67,20 |
| 70+ | 23 | 4 068 | 10 140 | 831 713 | <0,0001 | 53,63 | 30,10 | 69,23 |
| 18-59 | 81 | 20 655 | 61 542 | 3 879 966 | <0,0001 | **75,28** | 69,24 | 80,13 |
| 60+ | 45 | 7 667 | 20 654 | 1 689 920 | <0,0001 | **51,98** | 35,61 | 64,19 |

**Figure 2.**
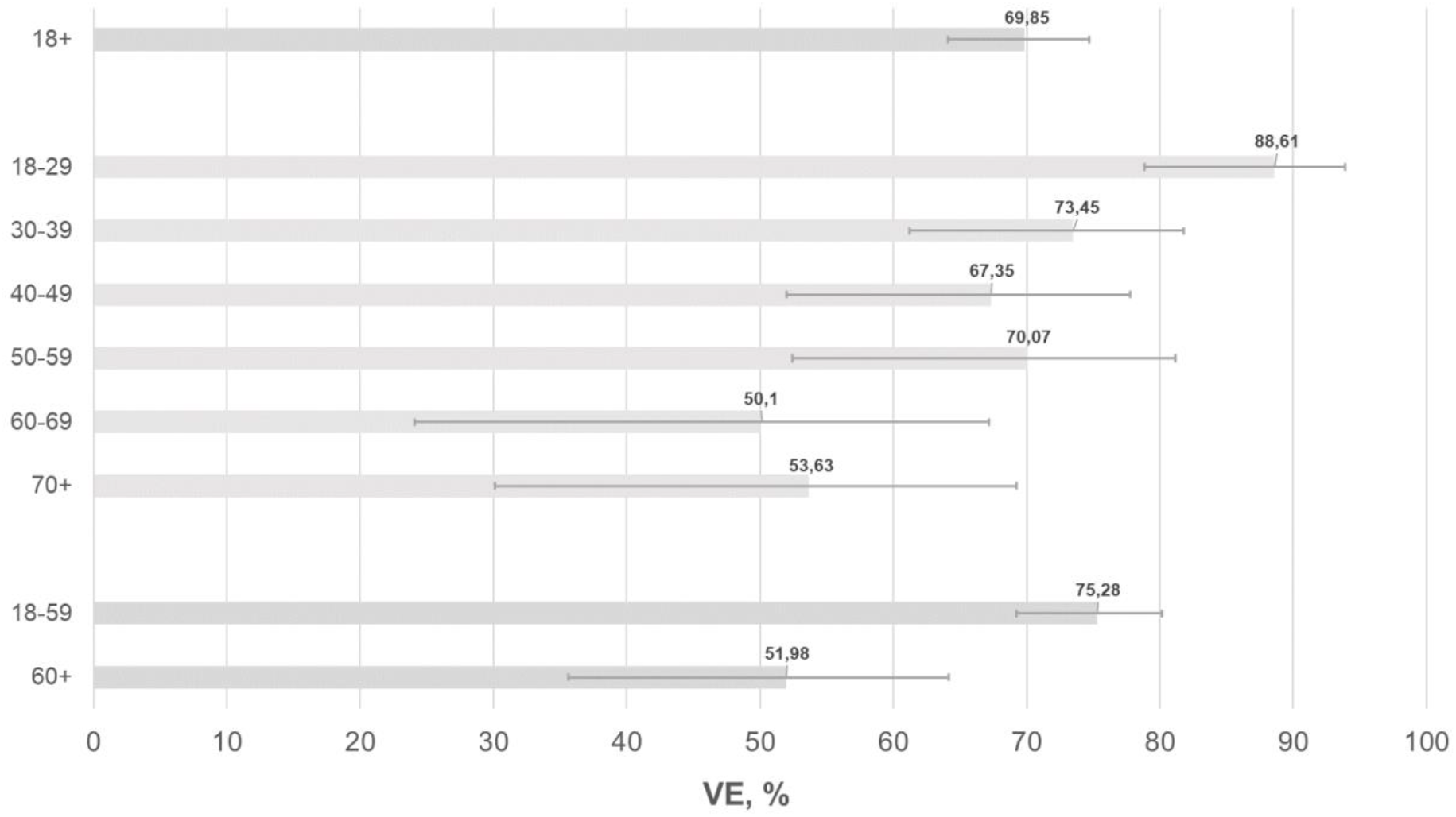
Efficacy of a single-dose vaccination in different groups. On the left, age groups are marked, columns indicate vaccine efficacy (VE, %) in age groups, whiskers show a 95% confidence interval.

## Discussion

It took about six months for the delta B.1.617.2 variant to displace other genetic lines in India [9]. At the same time, in India this variant rather quickly outstripped two other closely related lines B.1.617.1 and B.1.617.3. Our data show that in Moscow the displacement of endemic lines took no more than three months. It is obvious that the dynamics of the spread of the delta variant in different countries can be influenced by many factors. Meanwhile, today it is becoming clear that the dynamics of the spread of the delta variant little depends on the number of vaccinated in the country, and the main advantage of vaccination comes down to a decrease in the number of hospitalized, as well as deaths [5].

Several vaccine manufacturers have already announced the development of booster variants based on the S protein of delta variant [18-19]. It will take at least several months to confirm that the new vaccines are safe and more effective against the delta variant than the older vaccines. However, the decline in efficacy of the available vaccines may not be the same. Thus, there is reason to believe that the decrease in immunity over time in relation to the delta variant for the Astrazeneca drug may be less than for the Pfizer/BNT drug [20]. The difference in the dynamics of maintaining efficiency can be associated with both the characteristics of the vaccine platform and the structure of the antigen used. For example, BNT162b2 contains a biproline mutation to stabilize the S antigen in one conformation [21-22], which limits the formation of antibodies to a different conformation and, as a result, reduces the spectrum of neutralizing antibodies against new variants of coronavirus [23].

Sputnik V and Sputnik Light vaccines use a native S protein variant and an adenoviral delivery platform [11-12]. Our earlier data indicate a slight decrease in the neutralizing activity of the vaccinated sera against the delta variant [23]. At the same time, the clinical effectiveness of protection of the Sputnik V vaccine against delta remains at a level above 80% [13-14]. It should be noted that the assessment of the effectiveness of the Sputnik V vaccine by Barchuk and colleagues does not consider which vaccine the patient was vaccinated with. Evaluation of the effectiveness of the Sputnik V vaccine is made on the assumption that 96% of the vaccinated in the city were carried out using this vaccine. Meanwhile, it can be assumed that the proportion of those vaccinated among those who become ill after vaccination with other vaccines registered in Russia may not be the same.

There are few data on the effectiveness of the single-dose vaccine. Effectiveness of the Sputnik V first component in 60-79-years-old group in study in Argentina was 78.6% in the December 29, 2020 to March 21, 2021 period [24].

Here we made a calculations of the effectiveness of first component of Sputnik V vaccine using data from the Moscow registries of morbidity and vaccination against COVID-19. To calculate the efficiency, July 2021 was selected for which the total share of the delta B.1.617.2 variant and its descendants (advantage AY.12) was 99.5%. We calculated the size of a group that did not have preexisting immunity using a 54% non-immune rate, which was estimated thanks to constant serological monitoring carried out in Moscow. Our data indicate that the overall efficacy of a single-dose vaccine against the delta variant is 69.85% (95% CI, 64.08–74.70). This is significantly higher than for BNT162b2 and ChAdOx1 nCoV-19, for which the efficacy of a single dose against delta was estimated at 35.6% (22.7–46.4) and 30.0% (24.3–35.3), respectively [6]. Direct comparisons are difficult to make due to differences in study design. In the first case, a test-negative case-control design was used against the use in our case of an assessment based on the total upload of all data from the Moscow vaccination and morbidity registers with an adjustment for the size of the immune layer and calculation of the odds ratio (OR). Also, a significant difference may be that we took into account the effectiveness only within the first three months after vaccination. Evaluation of effectiveness during the first three months can be attributed to the limitation of our study. The choice of such an interval was caused by the fact that truly mass vaccination in Moscow began in April 2021, when the Moscow authorities began to tighten requirements for vaccination among people working in the service sector. Thus, many partially vaccinated people met dominant delta during summer 2021. Meanwhile, even for immunization with two doses, the revaccination period is set at six months after the primary immunization, so that the assessment of efficacy based on the first three months is an informative assessment.

Our results show that in age cohorts, the efficacy of a single-dose vaccine against the delta variant differs. Among young 18-29 years old, the effectiveness for the vaccine was 88.61% (95% CI, 78.81-93.88), in general among the 18-59 age group - 75.28 (95% CI, 69.24-80.13). However, with age, the effectiveness of the vaccine tended to decrease and in the group over 60 it was about 52%. Further studies are needed to determine the conditions for revaccination for patients who received the only one dose of the vaccine in different age cohorts.

## Conclusion

The results indicate a high efficacy of a single immunization with an adenovirus vaccine based on rAd26 (first component of Sputnik V vaccine) against SARS-CoV-2 delta variant among young and middle-aged people (18-59 yo), at least during the first 3 months after receiving the vaccine.

## Methods

### Study design and statistical analysis

We conducted an observational study of vaccine efficacy by screening method. In order to analyze the vaccine efficacy, it was necessary to select the right group of comparison. The most correct from our point of view is the use of the non-immune population for disease control in the analysis of the effectiveness of the vaccine. To calculate the immune layer, we used data obtained by the Moscow City Health Department, based on the results of continuous serological monitoring. For this monitoring, randomized groups of samples were formed among patients admitted routinely for treatment in “non-COVID” hospitals. The average number of patients examined is 10,000 per week. All patients underwent analysis of immunoglobulin G (IgG) in venous blood serum using reagents for the qualitative determination of IgG antibodies to SARS-CoV-2 by the immunochemiluminescence method on automatic CL series immunochemical analyzers manufactured by Mindray Medical International Limited (China). A total of 1,065,556 patients were examined from May 05, 2020 to June 25, 2021. The results of constant monitoring of Moscow make it possible to estimate the size of the immunological layer at the end of June 2021 was 46%. So, in order to count non-immune population, we took 54% of all population and set it as non-immune.

To assess the effectiveness of a single vaccination (first component Sputnik V), we used data on cases of COVID-19 among people who were vaccinated with only the first component of Sputnik V and did not receive a second dose of vaccine for various reasons. On September 6, 2021, data was downloaded on the number of cases among those vaccinated once, for whom the disease occurred in the interval between 21 days and 3 months after vaccination, and on the number of cases among the non-immune. We also downloaded data on the number of vaccinated once, for which the whole of July 2021 fell between 21 days and 3 months after vaccination, and on the number of non-immune at the end of June 2021. The effectiveness of vaccination was calculated in relation to the odds ratio using the formula: VE = (1-OR)*100.

Odds ratio (OR) and limits of 95% confidence interval of the odds ratio were calculated according to Tenny and Hoffman [25], p values were obtained by χ2 test or Fisher’s exact test.

The statistical analysis was done using Microsoft Excel, and GraphPad Prism v. 9.2.

### SARS-CoV-2 variant sequencing and analysis

The samples used in this study were collected as part of the ongoing surveillance of viral variability routinely conducted at the Gamaleya Center. Written informed consent was obtained from all subjects in accordance with the order of the Ministry of Health of the Russian Federation of July 21, 2015 N 474n. All samples were de-identified prior to receipt by the research team. The study was submitted to the Local Ethics Committee of the Gamaleya Center. The committee concluded that the study did not use identifiable biological samples and did not provide any confidential data. Consequently, according to the Local Ethics Committee rules and national regulations, this project does not require ethical approval.

Total RNA was extracted from patient’s nasopharyngeal swab’s and whole-genome amplification of SARS-CoV-2 virus genome was performed using ARTIC Network protocol as described previously [26-27]. Analysis of SARS-CoV-2 genetic diversity in Russia using GISAID was performed as described previously [28].

## Data Availability

All data produced in the present work are contained in the manuscript

## Acknowledgments

We would like to thank all the colleagues and all vaccinated people who took part in this study.

