## Supplementary Table S1 for "One-shot immunization with Sputnik Light (the first component of Sputnik V vaccine) is effective against SARS-CoV-2 Delta variant: efficacy data on the use of the vaccine in civil circulation in Moscow"

Supplementary table S1. Genetic SARS-CoV-2 lines identified during monitoring in Moscow in June-July 2021

|  |  | Lineage | | | | | | | |
| --- | --- | --- | --- | --- | --- | --- | --- | --- | --- |
|  |  | AT.1 | B.1.1.523 | B.1.1.525 | B.1.1.7 | B.1.351 | B.1.525 | AY.10 | AY.12 |
| June 2021 | Number | 2 | 32 | 0 | 4 | 2 | 3 | 0 | 450 |
|  | % | 0,31 | 4,89 | 0,00 | 0,61 | 0,31 | 0,46 | 0,00 | 68,70 |
| July 2021 | Number | 0 | 2 | 1 | 0 | 0 | 0 | 11 | 267 |
|  | % | 0,00 | 0,31 | 0,15 | 0,00 | 0,00 | 0,00 | 1,68 | 40,83 |
|  |  | AY.23 | AY.24 | AY.4 | AY.5 | AY.6 | B.1.617.2 | B.1.617.2 and AY | Total |
| June 2021 | Number | 0 | 0 | 4 | 1 | 0 | 157 | 612 | 655 |
|  | % | 0,00 | 0,00 | 0,61 | 0,15 | 0,00 | 23,97 | **93,44** |  |
| July 2021 | Number | 1 | 3 | 3 | 1 | 1 | 364 | 651 | 654 |
|  | % | 0,15 | 0,46 | 0,46 | 0,15 | 0,15 | 55,66 | **99,54** |  |
